# Protocol for a feasibility trial of a co-produced intervention: ‘Enabling Service Users with Severe Mental Illness to Learn about and Engage with Good Oral Health (SMILE)’

**DOI:** 10.64898/2026.10.05.26364521

**Authors:** MP Mishu, Natalia Kika, Emma Tkach, James Bird, Jade Cupac, Sarah Daniel, Michelle Horspool, Gordon Johnston, Helen Killaspy, Zoe Marshman, Louise Marston, Siyabonga Ndwandwe, Shengning Pan, Florence Watson, Emily Peckham

**Affiliations:** Department of Epidemiology and Public Health, University College London, London, United Kingdom; Sheffield Teaching Hospitals NHS Foundation Trust, Sheffield, United Kingdom; Research Development Unit, Sheffield Health and Social Care NHS Foundation Trust, Sheffield, United Kingdom; Research & Development Team, Tees, Esk and Wear Valleys NHS Foundation Trust, Darlington, United Kingdom; Independent Peer Researcher, Clackmannan, United Kingdom; Division of Psychiatry, University College London, London, United Kingdom; School of Clinical Dentistry, University of Sheffield, Sheffield, United Kingdom; Department of Primary Care and Population Health, University College London, London, United Kingdom; Department of Statistical Science, University College London, London, United Kingdom; College of Medicine and Health, Bangor University, Bangor, United Kingdom

**Author notes:** Corresponding author – Masuma Mishu.

**Keywords:** Feasibility trial, severe mental illness, oral health, co-production

## Abstract

**Background:** People with severe mental illness (SMI) report poorer oral health outcomes than the general population, are less likely to attend regular dental check-ups, and may require additional support to visit a dentist. Our previous research showed that interventions tailored to their needs and support from the mental health care system are needed to improve oral health outcomes in people with SMI. Although there is some provision of urgent dental care for people with SMI, there are currently limited available interventions that integrate dental and mental health care and provide dental support in a mental healthcare setting. To address this gap, the aims of the SMILE project are to 1) co-develop and 2) test the feasibility and acceptability of an oral health intervention for people with SMI, delivered by mental health professionals in secondary community mental health care settings. This paper describes the protocol for the feasibility and acceptability testing of the SMILE intervention.

**Methods:** This is a single-arm, non-randomised feasibility trial assessing the feasibility and acceptability of an intervention to support the oral health of mental healthcare service users with SMI. The intervention was co-produced with service users, carers, mental health and dental health professionals, and includes advice on maintaining good oral hygiene and lifestyle habits to maintain good oral health. The main aim of the study is to assess the feasibility and acceptability of delivering the intervention, using quantitative data collected at baseline, three and six-month follow-up, and qualitative interviews with participants and mental health staff involved in delivering the intervention.

**Results:** The study results will determine whether the intervention can be successfully delivered in secondary community mental health care settings and inform the most appropriate primary and secondary outcomes for a future trial.

**Conclusion:** The findings of this study will inform the feasibility of progression to the pilot stage of a randomised controlled trial to assess the clinical and cost effectiveness of delivering an oral health intervention in a mental healthcare setting.

**Trial registration:** This protocol was pre-registered via the ISRCTN (ref no.: ISRCTN44497673) on 19/02/2026.

## Background and rationale

Oral health among people with severe mental illness (SMI) is poorer than that of the general population. People with SMI had nearly three times the odds of having lost all their teeth than the general community (odds ratio = 2.81, 95% confidence interval = [1.73, 4.57])(1) and significantly higher decayed, missing, and filled teeth (mean difference = 5.0, 95% CI = 2.5-7.4) compared with the general community (2). They are less likely to visit a dentist for routine dental check-ups or treatment (3). These poor outcomes are due to several factors(4), including symptoms of mental illness that make people less motivated to brush their teeth regularly (5), high consumption of sugary food and drinks (6) and side effects of medications (e.g. dry mouth) (5). Lack of support to maintain oral hygiene and attend dental check-ups further exacerbates the problem^3^. Complications from untreated dental decay are common reasons for admission to hospital for physical health problems in this population (7) with associated resource implications for the health service. This group often requires specialist dental care and general anaesthesia, partly due to seeking treatment late. Appropriate support for preventing dental diseases and attending check-ups to identify problems earlier could significantly improve clinical and social outcomes, reduce dental and other associated health costs, and reduce the burden on dentists. However, to improve oral health, people with SMI require interventions tailored to their needs and that address the various barriers they face (Slack-Smith et al., 2017; Mishu et al., 2022). Beyond individual-level barriers such as mental health symptoms and mistrust in dental professionals, people with SMI also experience organisational barriers, for example, a lack of mental and dental health knowledge, staff capacity and resources. Finally, systemic barriers such as a lack of coordination and integration between dental and primary care and mental health professionals are usually attributed to a lack of clarity on which healthcare services hold the responsibility of supporting people with SMI with their oral health. Therefore, to address the multifaceted barriers people with SMI experience to receiving oral health support, an integrated and coordinated approach between mental health services and dental services is required(4). Currently, there are no such comprehensive interventions in place that not only target individual-level behaviour change but also work at a system level to integrate mental and dental healthcare services and aim to provide dental support in a mental healthcare setting.

Development of an acceptable and feasible complex intervention requires specific steps as outlined in the Medical Research Council guidance for the development of complex health interventions (8). Co-production with service users, clinicians, and researchers, ensures that interventions are accessible, relevant, and address the specific challenges experienced by people with SMI (9). The Enabling Service Users with Severe Mental Illness to Learn about and Engage with Good Oral Health (SMILE) is a co-produced intervention that will give advice and support on good oral health practices to people with SMI and be delivered by mental health professionals within routine mental healthcare. The SMILE project aims to address the lack of co-produced, integrated oral health interventions for people with SMI. The SMILE project follows two phases: 1) the development of an intervention to improve oral health in people with SMI, co-produced through a series of Patient and Public Involvement and Engagement (PPIE) activities and four co-production workshops with service users, carers, mental and dental health professionals; 2) the acceptability and feasibility testing of the co-produced intervention.

### SMILE Intervention

The SMILE oral health intervention was developed through a co-production process with service users, carers, dental health, and mental health professionals. We conducted four co-production workshops (two online, two in person), which were accompanied by concurrent PPIE meetings to gather input on the co-production process. Training resources for mental health staff delivering the intervention and an information leaflet for participants were also developed as part of this process.

The intervention will be delivered by NHS mental health staff (e.g., assistant psychologists, occupational therapists, physical healthcare nurses) who have received training from the study team in delivering the intervention. Training will be provided to the NHS mental health staff prior to delivering the intervention. A training manual and supporting materials (flip chart) will also be given to the mental health professionals to support their delivery of the intervention. As part of the intervention, participants will be given a leaflet which contains all of the intervention steps and advice to improve and maintain their oral health. Intervention delivery comprises one in person session lasting around 30 minutes and subsequent toothbrushing reminders for one month after the intervention. The intervention consists of three key components: 1) initiating a conversation about the importance of oral health for general wellbeing; 2) providing top tips/advice to maintain oral hygiene (toothbrushing, medication and dry mouth, sugary food and drink intake, tobacco and alcohol, regular dental check-ups); 3) sending four weekly toothbrushing reminders following the intervention delivery based on participants’ preferences.

Following the face-to-face intervention session, the interventionist will send a letter to the participant’s community mental health team to inform them about their participation in the study, and any concerns they have raised about their oral health. The community mental health team will also be encouraged to provide the participant with further support or signposting for accessing a dentist if they do not already have one.

PPIE contribution is an integral part of the study. We have established a PPIE group, specifically for this study, led by a person with lived experience and a mental health professional, supported by the research team. PPIE has been and will continue to be integrated into all stages of the study. The PPIE group consists of 10 members with lived experience of mental health problems, and mental and dental healthcare professionals. Those with lived experience represent a range of age, gender, ethnicity and mental health problems and experiences. The PPIE group met about every 4 months to discuss various components of the study, including the development of the intervention, staff training to deliver the intervention, potential outcome measures, participant information sheets, participant, recruitment, topic guides for the qualitative component. They will also contribute to data analysis and the dissemination of results.

This article describes our study protocol for Phase 2, the feasibility and acceptability testing of the co-developed SMILE intervention.

### Aims and Objectives

The main aim of this study is to assess the feasibility and acceptability of the co-produced SMILE intervention to improve oral health in people with SMI.

Objectives include:

1. To assess the feasibility and acceptability of the co-produced oral health intervention by measuring recruitment rate and uptake of and adherence to the intervention, and completion of outcome measures at follow-up points.
2. Assessing the relevance, appropriateness and completion of measures of oral health-related habits and oral health-related quality of life, and Economic evaluation questionnaires to be used for the main trial.
3. To explore the experiences of service users who receive the intervention and the professionals delivering the intervention.

## Methods

### Study Design

We will conduct a non-randomised, single-arm trial whereby all participants will be offered the intervention. The research team will collect quantitative data at baseline, three, and six months follow-up. We will also conduct qualitative interviews with approximately 10 intervention participants, four intervention delivery staff, and two research staff during the quantitative data collection period. This protocol is written according to adapted SPIRIT guidelines for feasibility trials (10), and in line with guidance on reporting the protocols of pilot and feasibility trials (11). The study was pre-registered via the ISRCTN (ref no.: ISRCTN44497673) on 19/02/2026.

### Study setting

We will conduct the study across two NHS mental health trusts in the North of England.

### Eligibility criteria

Inclusion criteria:

Service users will be eligible to join the study if they meet the following inclusion criteria and if they are currently receiving care from either of the participating NHS trusts:

1. Have a diagnosed severe mental illness (SMI): psychosis, schizophrenia, schizoaffective disorder, bipolar disorder or other non-organic psychosis (ICD codes to include are F20-29, F30-F31, F32.3, and F33.3).
2. Aged 18 years or above.
3. Comfortable communicating in English

Exclusion criteria:

Service users will not be eligible to take part in the study if they meet the following exclusion criteria:

1. Those in an acute psychiatric ward
2. Those who lack capacity to give informed consent to participate in the study, as guided by the Mental Capacity Act (MCA) 2005
3. Those who are not able to communicate in English

### How the objectives will be assessed

The main objective of the study is to assess the feasibility and acceptability of the SMILE intervention. This will be assessed by 1) reporting on rates of recruitment, intervention uptake, adherence, completion of follow-up measures and data missingness; 2) surveys completed by participants, and staff delivering the intervention to assess their perceptions of the feasibility and acceptability of the intervention quantitatively; 3) in-depth qualitative interviews with participants and staff delivering the intervention to explore their experiences of receiving or delivering the intervention.

We will explore the feasibility of a number of potential outcome measures for use in a future trial to assess the effectiveness of the intervention, including: a bespoke measure of participants’ oral health capturing oral health and dental access, tooth brushing habits and frequency, and dietary sugar intake; oral health-related quality of life (OHRQoL) measured using the Oral Health Impact Profile-14 (OHIP-14; (12)); health-related quality of life measured using EuroQol EQ-5D (13); and a bespoke questionnaire to assess the feasibility of collecting economic data from the participants.

### Sample size

As this is a feasibility study, there is no formal sample size estimate. There is limited guidance available regarding the optimum sample sizes for feasibility studies (14). Julious (15) recommends, as a rule of thumb, recruiting a minimum of 12 participants for a single-group feasibility study. We will exceed this and aim to recruit 30 participants.

No statistical methodology is required to control for a type 1 error since no formal hypothesis testing is being conducted. Potential outcomes are based on behavioural changes such as regular toothbrushing twice a day 6 months post-intervention. A previous study (16) suggested that around 50% of people with severe mental illness receiving standard care maintain a habit of regular toothbrushing. Considering an attrition rate of no more than 15% (17) at 6 months, and that we therefore retain 26 service users in the study, then our sample would be adequate to detect a regular toothbrushing rate of 30% to 70% with 95% confidence.

All analyses will be descriptive with mean (SD) or median (IQR) presented for continuous variables depending on distribution, and n/N (%) for categorical variables. We will provide 95% CI for retention, intervention uptake and adherence.

### Trial procedures

Potential participants will be identified using the following methods:

1. Screening of electronic health records (EHRs) and clinician caseloads
2. Research databases
3. Direct referrals
4. Advertising

Those who are interested will be provided with an information pack about the study and asked if they are willing to be contacted by a researcher about the study. Those who are willing will be contacted by a SMILE study researcher and given the opportunity to ask questions about the study before deciding whether they wish to take part. Written informed consent will be taken if they are happy to participate.

The participant identification and recruitment processes are described in more detail below.

### Identification of potential participants

#### 1. Screening

Researchers from the Trust’s research team will identify potentially eligible service users through EHR screening or caseload screening with clinicians. Anyone identified as eligible will be given or sent an information pack by the Trust’s research staff or their clinician, comprising an invitation letter, a participant information sheet and a consent to contact form. Interested service users can then either (i) give verbal consent to be contacted about the study to their care coordinator and ask their care coordinators to share their details with researchers either by email, phone, or verbally (ii) return the consent to contact form to researchers using a pre-paid envelope, (iii) contact researchers directly, using researcher contact details provided.

#### 2. Research databases

Where available, researchers based in the Trust’s research team will screen their Trust’s research database to identify potentially eligible participants. These databases identify service users who are willing to be contacted about relevant research, and consent to contact will therefore not be needed, as this has been previously provided. Researchers will then contact these service users to tell them about the study and ask if they are interested in hearing more. For those who are, the participant information pack will be provided, either via post or electronically depending on their preference, and the SMILE study researcher will then contact them and potentially recruit them into the study, gaining written informed consent as described below.

#### 3. Direct referral

Clinicians within the Trust can also refer eligible service users to the Trust’s research team and confirm that their patient is happy to be contacted about the study. Researchers will then send the information pack, as outlined above. Alternatively, clinicians will also be provided with study information packs to give eligible service users and then share participant contact details with the Trust research team who will contact the patient to discuss the study further.

#### 4. Advertising

Posters and flyers advertising the study will also be placed in community mental health services within the Trust and on the Trust’s social media pages. These will include contact details of the research team or service users can ask their clinician to contact the research team for them. If they meet eligibility criteria, they will then be provided with an information pack and recruited as before if willing.

### Consent

Once a potential participant has consented to their contact details being passed on, potential participants will be contacted by a SMILE researcher, based in the NHS Trust, inviting them to meet with a member of the SMILE researcher in the trust, the participant’s home, or another mutually convenient location. The potential participant will have a further opportunity to clarify any points they did not understand and ask any questions. A verbal explanation of the study will be given by the SMILE researcher, covering all the elements specified in the participant information sheet.

If the potential participant is still interested in taking part in the study, written informed consent will be taken. The participant must personally provide informed consent via the latest approved version of the Consent Form before any study specific procedures are performed. This consent may take place via signing a paper copy of the consent form, or via remote verbal consent taken over the phone or in an online meeting. Whichever method is used, participants will be asked to endorse every item on the consent form. They may do this verbally if remote or by initialling the relevant box on the paper consent form in person. If consent is taken remotely, this will be documented on a “Consent Form REMOTE”. Remote verbal consent will involve two-way communication in real time. The person who obtained the consent must be suitably qualified and experienced and have been authorised to do so by the Chief/Principal Investigator. A copy (physical or electronic) of the signed Consent form will be given to the participant. The original signed form will be retained at the study site.

Following signing the informed consent form, participants will complete the baseline questionnaire with a trained researcher. On completion of the baseline measures, participants will be offered the SMILE intervention. Intervention sessions will be arranged by an NHS mental health professional trained in the SMILE intervention who will liaise with the participant.

Participants’ flowchart in the study is shown in Figure 1.

**Figure 1.**
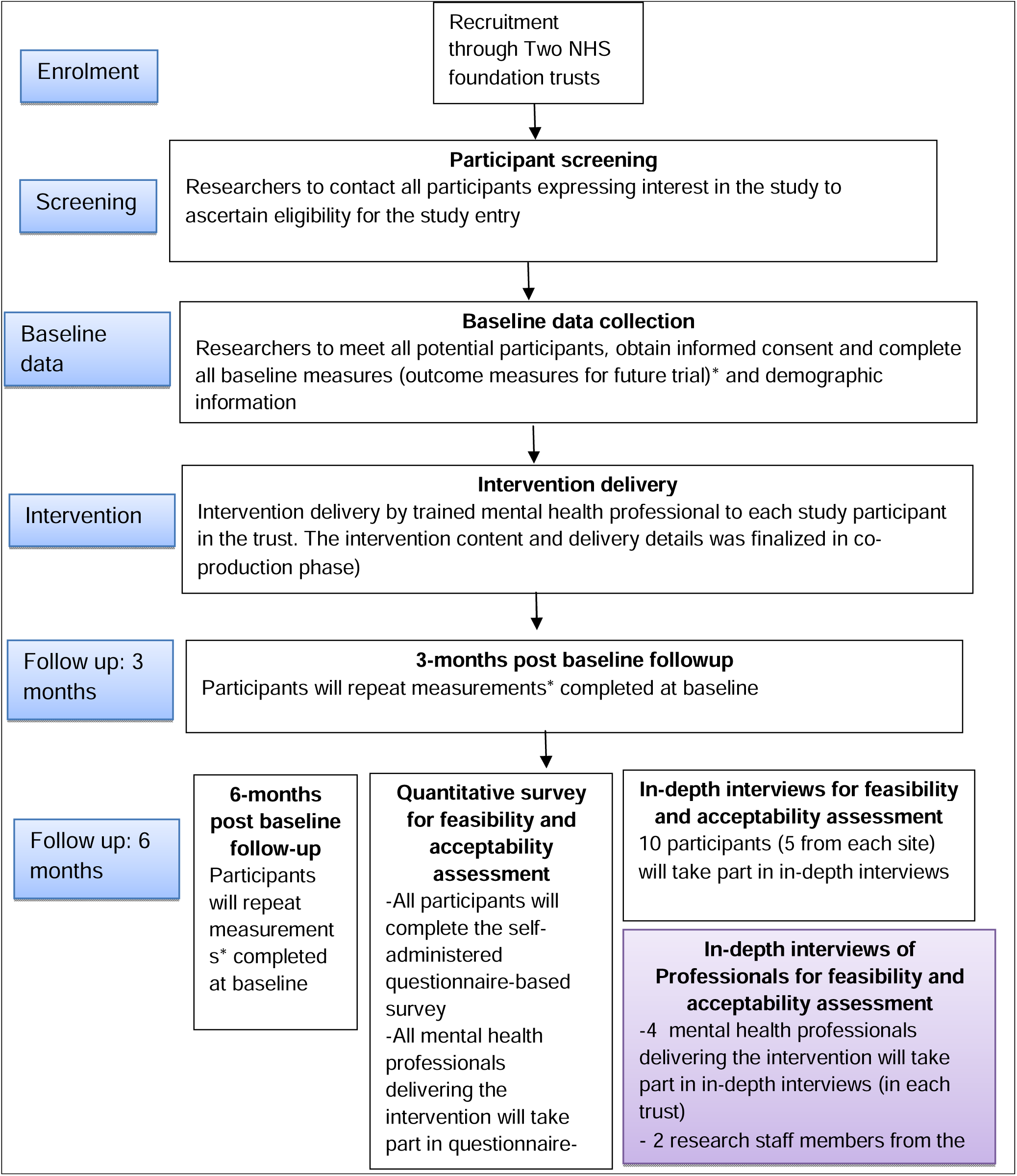
Flowchart of participants in the study.

#### Ineligible and non-consenting participants

All ineligible and non-consenting participants will be referred back to their care team. For ineligible participants, an explanation of the reasons why they are ineligible will be provided. A record will be kept by the research team of reasons for both ineligibility and non-consent.

### Data collection

We will invite participants to meet with a research assistant at baseline, 3-and 6-months after recruitment into the study to compete the self-report questionnaires. Participants can potentially complete the questionnaires over two separate appointments if needed.

After delivering the intervention to each participant, interventionists will complete a brief bespoke questionnaire report how long the intervention appointment took, and how it was delivered.

### Data management

Data management will be provided by the UCL Priment Clinical Trials Unit who adhere to their own Standard Operating Procedures (SOPs) relating to all aspects of data management, including data protection and archiving.

Study questionnaires will be finalised before the start of the trial. Collected data will be entered in to the REDCap database by the researchers as soon as possible after data collection with each participant.

All collection, storage, processing and disclosure of personal information will be performed in compliance with the Data Protection Act 2018. All investigators and study site staff will uphold the Act’s core principles. Any communications, reports or published results will not contain any personal data that could allow identification of individual participants.

### Data analysis

The acceptability of the intervention will be investigated using a mixed method approach from three data sources. We will analyse the quantitative and qualitative data separately and triangulate the results.

1. Recruitment, retention, fidelity to the intervention. Data on participant recruitment (number of eligible participants identified, approached and recruited), retention in the study, intervention adherence (attendance at the intervention session/s and contact with the participant regarding toothbrushing prompts), will be summarised descriptively using means and measures of spread for continuous data, and frequency and proportion of events for categorical data, using STATA 19 and R 4.5.0. A CONSORT diagram will be used to display the flow of participants through the study (draft flowchart is shown in Figure 1).

We will report intervention uptake (percentage of those who were eligible and who consented to the study), adherence (session attendance, number of participants who withdraw and the reasons for withdrawal), completion of follow-up measures (number of participants who completed follow-up questionnaires at 3 and 6 months from recruitment, including those who dropped out of the intervention). We will also summarise data missingness in the outcome questionnaires (number of items missing and which items received no responses). Fidelity to the intervention will be assessed using a simple checklist completed by the interventionist for each participant, on whether all content of the SMILE intervention was delivered in the face-to-face session.

2. Outcome measures: data will be analysed using descriptive statistics, in the same way as the recruitment, retention and fidelity data, reporting on completeness of the measures at baseline, 3 and 6 months follow-up.

3. Qualitative data. The qualitative data from our in-depth interviews will be analysed using the framework approach. These will explore the experience and perceptions about the study procedures, intervention content, and delivery from the perspectives of participants and interventionists, both using the APEASE framework (Acceptability, Practicability, Effectiveness, Affordability, Side-Effects, and Efficacy) (18). We will also assess the potential of the intervention-related training on capacity building in mental health staff who deliver the intervention.

### Progression criteria to the next stage of the research (full RCT)

The following criteria will inform whether results of this single-arm feasibility and acceptability study suggest progression to the pilot stage of a full scale randomised trial.

1. *Recruitment of participants:* Recruitment of ≥70% (i.e. an average of 2 participants per centre per month) being green (no further action needed), 50-<70% (1.5 – 2, less than 2 participants per month) being amber (minor modification is needed) and below 50% being red (major modification/ change is needed).
2. *Retention of participants:* Attrition of 15% at 6 months will be considered an acceptable limit. This attrition rate is based on a dental trial^12^ and three other NIHR funded large-scale pragmatic trials of behavioural interventions in people with SMI including, SCIMITAR+ (85% retention at 12 months).
3. *Intervention adherence:* 80-100% adherence will be considered acceptable
4. Results of the qualitative data analysis suggesting that participants and interventionists view the intervention as acceptable and feasible to deliver.

### Health economics analysis

We will estimate the intervention costs based on the delivery of the related activities over the intervention period and resources used by participants 6 months prior to the intervention and over the intervention period. We will also test the feasibility and acceptability of the bespoke health care and service use questionnaire. Analysis of the completeness of participants’ responses and use of listed services together with additional services recorded by participants, will enable further revision in order to collect most relevant data in a subsequent RCT and eliminate questions with few responses. The completeness and descriptive statistics of healthcare resources and health related quality of life measures (EQ-5D-5L and OHIP-14) will be summarised. No formal cost-effectiveness analysis will be undertaken as the feasibility study is not sufficiently powered, and there is no comparator/control arm assessed. In addition, the interventionists will complete questions on appointment duration and delivery format.

### Safety and adverse events

Definition of Adverse event (AE): This is any untoward medical occurrence in a participant to whom an intervention has been administered, that may be related to that intervention. Definition of Serious adverse event (SAE): Any untoward medical occurrence that results in death; is life threatening; requires in-patient hospitalisation or prolongation of existing hospitalisation; results in persistent or significant disability or incapacity; or consists of a congenital anomaly or birth defect. Other ‘important medical events’ may also be considered serious if they jeopardise the participant or require an intervention to prevent one of the aforementioned consequences. A planned hospitalisation for a pre-existing condition is not considered an SAE.

#### Which adverse events will be recorded?

This study is considered a low-risk research study. We do not expect any AE or SAE due to the intervention (oral health related advice and related support). However, we will record and report all AEs and SAEs during the 6 months after receiving the intervention that are deemed to be related to taking part in the trial or intervention. All SUSAR events will be reported to and the REC by UCL Priment clinical trials unit. We will also record all deaths, regardless of causality. All (S)AEs will be recorded by SMILE researchers on a study-specific case report form and reviewed by an independent clinician/mental health specialist at the NHS site.

### Safety measures and risk management

Safety measures have been built into the intervention to ensure participant safety, such as suitably experienced and qualified health professionals delivering the intervention and appropriate procedures to follow if a participant demonstrates safety concerns or distress.

If a participant discloses information that suggests that they or others are at serious risk of harm, the research team will follow good clinical practice and follow a study risk protocol for the reporting/monitoring of risk during all encounters with study participants. The research team has a wealth of experience in developing and implementing risk protocols for use in studies involving people with SMI.

Distress management - If a participant expresses any signs or reports any distress during any study related activity (e.g. meeting with a member of the research team or during intervention delivery) study specific guidance will be used to guide the researcher or interventionist for the management of this situation. Trust based researchers will also follow Trust policies for the management and reporting of risk. Incidences of low level distress will be reviewed during scheduled supervision meetings. If a participant is referred to the local mental health team for management of their distress the Chief Investigator (or delegated research clinician i.e. local PI) will be informed within 24 hours. Participants will be reminded at each phase of the study of their right to withdraw. In addition, consent will be checked throughout the study phases and during data collection, which will be conducted in a manner to minimise the potential distress or inconvenience to participants.

### Ethics

Research Ethics Committee approval has been obtained from the NHS West Midlands - South Birmingham Research Ethics Committee (reference number: 361009).

### Confidentiality

All data collected from participants during the feasibility study will be confidential and will not contain any information that may lead to the identification of an individual. All participants will be assigned with an ID number which will be used on any questionnaires they complete to maintain anonymity.

### Data storage

All information will be stored securely and adhere to GDPR regulations and the principles of the Data Protection Act (2018).

### Dissemination plans

The long-term aim of the intervention is to improve oral health in people with SMI.

This research will produce four key short-term outputs: 1) a prototype ‘SMILE with good oral health’ intervention package; 2) training resources for interventionists; 3) Co-produced roadmap for a service-level planning to be able to integrate and deliver the intervention in routine care; 4) Peer reviewed articles, articles for third sector organisations and websites, and conference presentations.

## Discussion

This paper outlines the protocol for the feasibility and acceptability study of SMILE; a novel, intervention to improve oral health of people with SMI, a population whose oral wellbeing has historically been systemically underserviced, and poorer compared to the general population (4). The SMILE intervention depends on an educational approach to improving oral health amongst people with SMI, an approach which has been deemed feasible, acceptable and effective in an evaluation of a similar intervention created in the United States (19). This intervention was a health educational program, which included oral health educational materials, meant for people with SMI, and implemented and tested in one community centre, and three drop-in psychiatric centres. evaluation found this intervention was appropriate as an educational program for improving oral health in those with SMI. However, the SMILE intervention differs from the oral health education program developed by Lapidos et al. (19) as it aims for integration within the mental healthcare system.

Developing this kind of system-level oral health intervention, and assessing its feasibility and acceptability, required co-production processes with stakeholders who are central to the success of this intervention: people with SMI who use mental health services, as well as mental health and dental health professionals. This co-production approach relied upon PPIE methods, which have been successful in the development of other system-level health interventions (8).

The outcomes of SMILE’s feasibility and acceptability will guide future research and impact in this growing field at the intersection of oral and mental health. This intervention represents an innovative approach to improving oral health equity. If SMILE is deemed feasible and acceptable, we intend to progress to the next stage of its assessment, a pilot trial, prior to a full-scale RCT that will evaluate its efficacy and cost-effectiveness.

## Declarations

## Ethics approval and consent to participate

Research Ethics Committee approval has been obtained from the NHS West Midlands - South Birmingham Research Ethics Committee (reference number: 361009). Informed consent will be obtained from all participant prior to data collection.

## Consent for publication

Not applicable

## Availability of data and materials

Not applicable

## Competing interests

The authors declare that they have no competing interests.

## Funding

This project is funded by the National Institute for Health and Care Research (NIHR) under its Research for Patient Benefit (RfPB) Programme (Grant Reference Number NIHR206746/PG-PB-XXXX-XXXXX). The views expressed are those of the author(s) and not necessarily those of the NIHR or the Department of Health and Social Care.

## Authors’ contributions

M.P.M, N.K., E.T., H.K., and E.P. wrote the original manuscript draft. M.P.M. and E.P. conceptualised and acquired funding for the study. All authors reviewed and edited the manuscript draft.

## Data Availability

Not applicable (study protocol)

## Acknowledgements

Not applicable.

## Notes

### Competing Interest Statement

The authors have declared no competing interest.

### Clinical Trial

ISRCTN44497673

